# Sensitivity of rapid antigen tests for COVID-19 during the Omicron variant outbreak

**DOI:** 10.1101/2022.06.13.22276325

**Authors:** Michio Murakami, Hitoshi Sato, Tomoko Irie, Masashi Kamo, Wataru Naito, Tetsuo Yasutaka, Seiya Imoto

## Abstract

**Background:** Rapid antigen tests have been used to prevent the spread of the coronavirus disease 2019 (COVID-19); however, there have been concerns about their decreased sensitivity to the Omicron variant.

**Aims:** In this study, we compared the sensitivity and specificity of the rapid antigen and the polymerase chain reaction (PCR) tests among the players and staff members of the Japan Professional Football League and clubs. Furthermore, we evaluated the relationship between the sensitivity and the duration from the onset of the symptoms to testing, the manufacturer of the rapid antigen test kits, and the PCR test analyte.

**Design and methods:** This was a retrospective observational study. We used 656 results from both the rapid antigen and PCR tests for COVID-19 using the analytes collected on the same day from January 12 to March 2, 2022, during the Omicron variant outbreak in Japan.

**Results:** The sensitivity of the rapid antigen test compared with the PCR test was 0.63 (95% confidence interval: 0.54–0.72) and the specificity was 0.998 (95% confidence interval: 0.995–1.000). There were no significant associations between the sensitivity and the duration from the onset of the symptoms to testing (including asymptomatic cases in the category), vaccination status, manufacturer of the rapid antigen test kit or PCR analyte (*P* > 0.05) with small effect sizes (Cramer’s V or φ: ≤ 0.22).

**Conclusions:** Even during the Omicron outbreak, the sensitivity of the rapid antigen tests did not depend on the duration from the onset of the symptoms to testing.

## Introduction

To prevent the spread of the coronavirus disease 2019 (COVID-19), active testing has been used to identify and isolate infected individuals, especially in populations at high risk of infections ^1^. Among the various testing methods including the reverse transcription-polymerase chain reaction (PCR) test, antigen quantitative test, and rapid antigen test, the rapid antigen test is less sensitive, but it has the advantage of being inexpensive and providing prompt test results ^2^. In particular, highly-frequent routine testing using rapid antigen test kits is more promising in reducing the spread of infection than highly-sensitive, but low-frequent testing ^3^. It has been noted; however, that the sensitivity of the rapid antigen tests may be lower in Omicron variants than in previous variants ^4,5^. In addition, the sensitivity of the rapid antigen tests may be particularly lower during the few days after infection (preprint)^6^. Since the testing and identification of infected individuals is more effective in controlling the spread of infection during the short period between infection and testing, there is concern that the lower sensitivity of the rapid antigen test during the short period after infection, may reduce the effectiveness of the testing system in the population. However, contrary to this, a previous study reported no large differences in the analytical sensitivity of the rapid antigen test in a comparison between representative Delta and Omicron isolates, using ten test kits ^7^. In another case study with human participants, there was also no difference in the sensitivity of the rapid antigen test between the Delta and Omicron variants (preprint) ^8^. Since both rapid antigen tests and other tests (e.g. PCR tests) must be performed using the analytes collected on the same day from the same individuals to evaluate the sensitivity of the rapid antigen tests, studies based on human participants have been limited ^9^ and these findings were not sufficient.

The Japan Professional Football League, a professional league of the most popular sports in Japan, collected the results of rapid antigen and PCR tests for COVID-19 among players and staff members in order to maintain and promote its activities ^10^. Since January 2022, rapid antigen tests were conducted twice a week on a regular basis, and moreover, additional PCR tests were often conducted on players and staff members in the clubs where infected individuals were identified. Consequently, from January 12 to March 2, 2022, during the period when the Omicron variants emerged in Japan, the number of cases in which both rapid antigen and PCR tests were performed on the same day exceeded 650, which made it possible to evaluate the sensitivity of the rapid antigen and PCR tests.

In this study, we compared the results between the rapid antigen and PCR tests for COVID-19 among the players and staff of the Japan Professional Football League and clubs to determine the sensitivity and specificity of the rapid antigen and PCR tests. We then assessed the relationships between the sensitivity and the duration from the onset of the symptoms to testing, the manufacturers of the rapid antigen test kit, or the analytes of the PCR tests.

## Methods

### Ethics

This study was conducted with the approval of the Ethics Review Committee of the Institute of Medical Science, University of Tokyo (approval number 2022-1-0421). Testing was not conducted originally for research purposes and the Japan Professional Football League does not have personal information on all the results. Therefore, information about this study was disclosed on the websites of the Institute of Medical Science of the University of Tokyo and the Japan Professional Football League to provide participants with the opportunity to opt out of the study. The person in charge of each club also provided information about the study to potential participants (players and staff members).

### Participants

This study was a retrospective observational study. We obtained the test results from January 12, to March 2, 2022. This was the period of the Omicron variant outbreaks in Japan (98.92% on February 7, 2022) ^11^. The data included a total of 656 cases in which both rapid antigen and PCR tests were performed using the analytes collected on the same date from players and staff members of the Japan Professional Football League and clubs. In the process of collecting the test results from players and staff members, some of the cases in which both tests were negative may not have been available: i.e., the number of cases reported in this study in which both tests were negative may have been smaller than the actual number.

### Survey items

The information used in this study included the positivity or negativity of each test, the presence or absence of symptoms, duration between the onset of symptoms and testing, vaccination status (i.e., whether the participants were vaccinated: at least once, none, or unknown), the manufacturer of the rapid antigen test kit, the analyte of the PCR test, and the type of test (“regular test,” defined by the use of a routine rapid antigen test twice a week by the Japan Professional Football League or a “voluntary test” other than a routine test). The onset of symptoms was based on the tally by the Japan Professional Football League, which comprised the individuals’ self-reported information that their health condition was different from usual (e.g., fever, sore throat). The date of the onset of symptoms represented the date when the symptom developed. Asymptomatic cases represented those who did not exhibited symptoms up to the time of testing and after.

The rapid antigen test was performed using nasal swab samples, and the kits were the Abbott Panbio™ COVID-19 Antigen Rapid Test or the Roche SARS-CoV-2 Rapid Antigen Test. The analytes for the PCR test were saliva or a nasal swab. Both analytes were collected by the participants themselves, the testing managers, or physicians. The players and staff members of the Japan Professional Football League and the clubs received lectures from their physicians on how to collect samples. Each club sent their analytes to a medical or measuring laboratory for PCR testing.

A Ct (threshold cycle) value of < 40 was considered as positive. Since information on the manufacturer of the rapid antigen test kits and the PCR analytes was not available on an individual basis, we instead matched the individuals and their club using the information that was obtained from a survey of how each club conducted testing during the period. The clubs determined whether the manufacturer of the rapid antigen test kit was Abbott, Roche, or either, and whether the PCR analytes were saliva, nasal swab, either, or other. The results (positivity or negativity) of the rapid antigen test among each of the 103 PCR-positive cases according to the duration from the onset of the symptoms to testing (including asymptomatic cases in the category) were reported on the website of the Japan Professional Football League ^12^.

### Statistical analysis

In this study, the sensitivity and specificity of the rapid antigen test against PCR test were first calculated by comparing the results (positivity or negativity) between both tests. Next, among the cases with positive PCR results, the chi-square test or Fisher’s exact test was performed to investigate the associations between the results of the rapid antigen test (positivity or negativity) and the duration from the onset of the symptoms to testing (including asymptomatic cases in the category), vaccination status, manufacturer of the rapid antigen test kit, PCR analyte, or test type. As an additional stratified analysis, only vaccinated individuals, those whose rapid antigen test kit manufacturer was Abbott, and those whose PCR analyte was saliva were used to examine the relationships between the rapid antigen test result (positivity or negativity) and the duration from the onset of the symptoms to testing (in categories asymptomatic included) using the chi-square test or Fisher’s exact test. In this stratified analysis, -2 and -1 days were grouped together as one category for the duration from the onset of the symptoms to testing. Similarly, one and two days were combined into one category.

IBM SPSS version 28 and R 4.2.0 ^13^ were used for the statistical analysis.

## Results

Of the 656 cases, 65 were positive for both the rapid antigen and PCR tests, 38 negative for the antigen tests and positive for the PCR test, one was positive for the rapid antigen test and negative for the PCR test, and 552 were negative for both (Table 1). The sensitivity of the rapid antigen tests compared with the PCR tests was 0.63 (95% confidence interval (CI): 0.54–0.72) and the specificity was 0.998 (95% CI: 0.995–1.000).

**Table 1.** Results of the rapid antigen and polymerase chain reaction (PCR) tests.

|  |  | PCR |  |  |
| --- | --- | --- | --- | --- |
|  |  | + | – | Total |
| Rapid<br>antigen | + | 65 | 1 | 66 |
|  | – | 38 | 552<sup>a</sup> | 590 |
|  | Total | 103 | 553 | 656 |

<sup>a</sup> The values of the number of participants with both negative rapid antigen and PCR tests shown in the table may be smaller than the actual values.

For the 103 cases that were positive for the PCR test, there were no significant associations between the sensitivity and the duration from the onset of the symptoms to testing (Cramer’s V = 0.146, *P* = 0.837; Table 2). Similarly, the sensitivity was not associated significantly with the vaccination status, kit manufacturer, PCR analyte, or test type (in the order: Cramer’s V = 0.220, *P* = 0.073; Cramer’s V = 0.204; *P* = 0.118; Cramer’s V = 0.217, *P* = 0.108; φ = 0.012, *P* = 0.904; Table 3). Among those whose PCR analyte was saliva (n = 80), the sensitivity was 0.58 (95% CI: 0.47–0.68).

**Table 2.** Associations between the sensitivity of the rapid antigen tests compared with the polymerase chain reaction (PCR) tests and the duration from the onset of the symptoms to testing, vaccination status, kit manufacturer, PCR analyte, or test type.

| Items | | Rapid<br>antigen: +<br>PCR: + | Rapid<br>antigen: –<br>PCR: + | Sensitivity | $\phi$ or<br>Cramer's V | P |
| --- | --- | --- | --- | --- | --- | --- |
| Duration from<br>the onset of<br>the<br>symptoms to<br>testing | –2 days | 3 | 1 | 0.75 | 0.146 | 0.837<sup>a</sup> |
|  | –1 day | 5 | 3 | 0.63 |  |  |
|  | 0 day | 20 | 16 | 0.56 |  |  |
|  | 1 day | 12 | 5 | 0.71 |  |  |
|  | 2 days | 5 | 4 | 0.56 |  |  |
|  | Asymptomatic | 20 | 9 | 0.69 |  |  |
| Vaccination | Yes | 43 | 27 | 0.61 | 0.220 | 0.073<sup>a</sup> |
|  | No | 9 | 9 | 0.50 |  |  |
|  | Unknown | 13 | 2 | 0.87 |  |  |
| Kit<br>manufacturer | Abbott | 33 | 12 | 0.73 | 0.204 | 0.118<sup>b</sup> |
|  | Roche | 8 | 9 | 0.47 |  |  |
|  | Either | 24 | 17 | 0.59 |  |  |
| PCR analyte | Saliva | 46 | 34 | 0.58 | 0.217 | 0.108<sup>a</sup> |
|  | Nasal swab | 9 | 2 | 0.82 |  |  |
|  | Either or other | 10 | 2 | 0.83 |  |  |
| Test type | Regular | 23 | 13 | 0.64 | 0.012 | 0.904<sup>b</sup> |
|  | Voluntary | 42 | 25 | 0.63 |  |  |

<sup>a</sup> Fisher's exact test. <sup>b</sup> Chi-square test.

**Table 3.** Associations between the sensitivity of the rapid antigen tests compared with the polymerase chain reaction (PCR) tests and the duration from the onset of the symptoms to testing: a stratified analysis.

|  | Rapid<br>antigen: +<br>PCR: + | Rapid<br>antigen: –<br>PCR: + | Sensitivity | Cramer's V | *P* |
| --- | --- | --- | --- | --- | --- |
| Vaccine: yes (n=70) |  |  |  |  |  |
| –2 days or –1 day | 7 | 3 | 0.70 | 0.084 | 0.955<sup>a</sup> |
| 0 day | 15 | 11 | 0.58 |  |  |
| 1 day or 2 days | 7 | 4 | 0.64 |  |  |
| Asymptomatic | 14 | 9 | 0.61 |  |  |
| Kit manufacturer: Abbott (n=45) |  |  |  |  |  |
| –2 days or –1 day | 4 | 3 | 0.57 | 0.181 | 0.688<sup>a</sup> |
| 0 day | 13 | 3 | 0.81 |  |  |
| 1 day or 2 days | 3 | 1 | 0.75 |  |  |
| Asymptomatic | 13 | 5 | 0.72 |  |  |
| PCR analyte: saliva (n=80) |  |  |  |  |  |
| –2 days or –1 day | 6 | 4 | 0.60 | 0.087 | 0.895<sup>b</sup> |
| 0 day | 16 | 14 | 0.53 |  |  |
| 1 day or 2 days | 10 | 8 | 0.56 |  |  |
| Asymptomatic | 14 | 8 | 0.64 |  |  |

<sup>a</sup> Fisher's exact test. <sup>b</sup> Chi-square test.

A stratified analysis of 70 vaccinated individuals showed no significant association between the sensitivity and the duration from the onset of the symptoms to testing (Cramer’s V = 0.084, *P* = 0.955). Similarly, the stratified analysis of 45 individuals whose used Abbott and of 80 individuals whose PCR analytes was saliva showed no significant associations between the two (in the order: Cramer’s V = 0.181, *P* = 0.688; Cramer’s V = 0.087, *P* = 0.895).

## Discussion

In this study, using 656 cases, we compared the rapid antigen and PCR tests for COVID-19, that were conducted on the same day among players and staff members of the Japan Professional Football League and clubs from January to March 2022, when the Omicron variant emerged, in order to determine the sensitivity and specificity of the rapid antigen test against the PCR test. We also investigated on the relationship between the sensitivity and the duration from the onset of the symptoms to testing, vaccination status, rapid antigen test kit manufacturer, PCR analyte, or test type.

The sensitivity was 0.63 (95% CI: 0.54–0.72) and specificity was 0.998 (95% CI: 0.995–1.000). The specificity was possibly an underestimate because there may have been fewer reports on the number of cases that were negative for both tests than the actual number. The sensitivity was not associated significantly with the duration from the onset of the symptoms to testing. Consistent results were found in the stratified analysis of only those who were vaccinated, those whose kit manufacturer was Abbott, and those whose PCR analyte was saliva. Overall, the effect sizes were small (Cramer’s V < 0.2). Furthermore, the sensitivity was associated insignificantly with vaccination status, kit manufacturer, PCR analyte, or test type (Cramer’s V or φ ≤ 0.22).

The results obtained in this study indicated that the sensitivity of the rapid antigen test compared to the results of the PCR test was independent of the duration from infection to testing or the presence or absence of symptom onset. This result was contrast to that of the previous report (preprint) ^6^: sensitivity of the rapid antigen test (Abbott or Quidel) compared with that of the PCR test (analyte: saliva) was 0.25 within two days from the first positive PCR test to the target testing and 0.9 since three days. The sensitivity in this study was higher than the sensitivity of the previous study (i.e., 0.25 within two days from the first positive PCR test to the target testing). The reason for this difference was not clear. One possible explanation is that the players and staff members who were the participants of this study received lectures from their physicians on how to collect analytes and that the tests were performed routinely, so that the analytes were collected appropriately. The sensitivity of the rapid antigen tests may decrease when the tests are not performed according to the manufacturers’ instructions for use ^14^. Proper analytes collection can lead to a high sensitivity.

The results of this study, which showed that the sensitivity of the rapid antigen test compared with the PCR test was 0.63 (95% CI: 0.54–0.72), may be used in combination with a model analysis to provide the fundamental knowledge required to establish a highly effective and efficient testing system. For example, a model analysis has estimated that the use of frequent rapid antigen testing is more effective than infrequent PCR testing in reducing the infection risk among populations such as professional sport players and staff members ^15^. Under the assumption of an incubation period of five days and an R_0_ of 4, the infection risk (defined as “number of infected individuals remaining at the end of the two-week isolation”) among population, in which a daily rapid antigen test with a sensitivity compared with a PCR test of 0.6 that was conducted for two weeks, was estimated to be as effective as when PCR testing was performed every three days ^15^. Similarly, the sensitivity of 0.5 and 0.7 was equivalent to a PCR test being performed once every four days and every two days, respectively. Since the cost of the rapid antigen test is approximately 1/10 that of the PCR test, the rapid antigen test can be performed more frequently than the PCR test under the same financial resources, and is therefore expected to be highly effective in controlling infection. However, since the Omicron variant is more infectious than previous variants ^16^ and has a shorter incubation period ^17^, future testing strategies are expected to be combined with further model evaluations to match the characteristics of the Omicron variant.

This study had some limitations. First, the manufacturer of the test kits and the analytes used in the PCR tests were based on the data provided by the clubs, and it was not possible to identify the manufacturer or analytes of some participants. In this study, however, we found that there were no significant differences in the sensitivity by including the group that could not identify the manufacturer or analyte as a separate category in the analysis. We also confirmed that there was no association between the sensitivity and the duration from the onset of the symptoms to testing by performing a stratified analysis of only those for whom the manufacturer was Abbott or the PCR analyte was saliva. Second, this study did not provide clinical diagnostic information on COVID-19. Therefore, it was not possible to assess the sensitivity of the rapid antigen test against the clinical diagnosis. Third, we could not obtain information on the participants’ age, gender, presence or absence of underlying diseases, and history of COVID-19 infection. The Ct values for the PCR tests were also only available from some of the participants. Therefore, it was not possible to evaluate the association between the sensitivity of these items. Fourth, the participants of this study were professional sport players and staff members and are therefore considered, in general, to be a healthy population. Cautions are therefore required in applying the findings of this study in populations with different characteristics, such as children, elderly, and those with underlying diseases.

Despite such limitations, this study analyzed the sensitivity and specificity of the rapid antigen test against the PCR test during the Omicron variant outbreak, and found that the sensitivity was independent of the duration from the onset of the symptoms to testing.

## Supporting information

STROBE

## Data Availability

We have included all the data produced in the present work in the manuscript. Note that the raw data used in the study were provided by Japan Professional Football League, as described in this paper. We are unable to attach all the raw data for each participant in this paper due to the ethical restrictions.

## Acknowledgements

We would like to thank Editage (www.editage.com) for English language editing.

## Funds

No external funds were used in this study.

